# SARS-Cov-2 Viral Load as an Indicator for COVID-19 Patients’ Hospital Stay

**DOI:** 10.1101/2020.11.04.20226365

**Authors:** Salman Al Ali, AbdulKarim AbdulRahman, Omar Yaghi, Essam M. Janahi, Manaf Al-Qahtani

## Abstract

**Background/objective:** The novel coronavirus disease 2019 (COVID-19) pandemic poses a global threat to the public health. There is a challenge in measuring the patient’s length of hospital stay and managing the healthcare resources to handle the situation successfully. Our objective is to use the qPCR cycle of threshold (Ct) as a tool in evaluating the severity of the infection and hence the length of hospital stay to better utilize and manage the healthcare resources.

**Methods:** This cross sectional study was carried out on 306 patients who admitted to COVID-19 care centers in Kingdom of Bahrain from 20^th^ March 2020 to 5^th^ April 2020. Standard qPCR was used to estimate the viral load and data were analyzed to investigate the relationship between Ct values and various variables.

**Results:** Out of 306 patients, 2 deaths, 1 active stable case and 303 recovered cases were reported. Ct value was significantly and negatively associated (P value <0.001) with length of hospital stay. The viral clearance was also inversely associated with the Ct values.

**Conclusion:** Ct value was inversely associated with hospital stay duration (and time to viral clearance), higher the Ct value is indicative of faster time to viral clearance. This association could help to better manage the infection and resources allocation.

## Introduction

Coronavirus disease 2019 (COVID-19) is caused by the newly identified strain of the Coronavirus family Severe Acute Respiratory Syndrome Coronavirus 2 (SARS-CoV-2) ^1^. Whilst some patients may have mild symptoms, others develop a severe respiratory distress syndrome with features of a blood coagulopathy requiring intensive support ^2^. As a result, COVID-19 has rapidly evolved into a pandemic that has put a severe burden on the healthcare services of some countries, notably beds for both hospitalization and intensive care facilities ^3^. Testing aids case identification, case isolation and contact tracing, all of which are vital to limit coronavirus spread ^4^. Currently, the main diagnostic test for SARS-CoV-2 is the real-time Reverse Transcription Polymerase Chain Reaction (RT-PCR) ^5^.The RT-PCR result is influenced by the Cycle Threshold (Ct-Value) that has been suggested as a quantitative test to indirectly assess the viral load ^6^ with severe cases having a much higher viral load than mild cases ^7^. Therefore, the aim of this study was to measure the viral load determined by the Ct value and relate that to viral clearance to determine if this could predict length of hospital stay, as that may help guide healthcare resource allocation.

## Methods

### Patients and Sample Collection

A cross sectional study was undertaken on 306 patients who were admitted to COVID-19 care centers in the Kingdom of Bahrain between March 20^th^, 2020 and April 5^th^, 2020. Of those 306 patients, 303 had recovered from their COVID-19 disease. Data were recorded for demographics, Ct-value on admission, time to viral clearance, and data on the length of stay and patient outcomes. At the time of the study, the local protocol in Bahrain admits all positive cases to treatment/isolation facilities regardless of being symptomatic or asymptomatic. The patients are discharged when they have two negative nasopharyngeal and oropharyngeal swabs on two consecutive days. All admitted patients are tested after three days of being asymptomatic and, if negative, the test is repeated after 24 hours to confirm viral clearance, following which the patients are discharged. Therefore, the length of stay was the same as the time to viral clearance and the minimum stay of patients is four days.

### RNA Extraction and SARS-Cov-2 RNA Amplification

The diagnostic test used for SARS-CoV-2 was performed using qPCR on nasopharyngeal and oropharyngeal samples. The samples were transferred to a viral transport media immediately after collection and transported to a COVID-19 laboratory for testing. Diagnosed cases were a heterogeneous sample of tested patients, they included symptomatic and asymptomatic individuals with recent exposure to a confirmed COVID19 case or travelers arriving from high risk countries. PCR test was conducted using Thermo Fisher Scientific (Waltham, MA) TaqPath 1-Step RT-qPCR Master Mix, CG on the Applied Biosystems (Foster City, CA) 7500 Fast Dx RealTime PCR Instrument. The assay used followed the WHO protocol from Charité Virology, Berlin, Germany ^8^. and targeted the E gene. If the E gene was detected, the sample was then confirmed by RdRP and N genes. The E gene CT value was reported and used in this study. CT Values >40 were considered negative. Positive and negative controls were included for quality control purposes.

### RT-qPCR Analytical Validation

Linear range was determined using serial log_10_ dilutions of standard RNA and was established between 3·5 × 10^3^ to 3·5 × 10^12^ copies of RNA/mL, with amplification efficiency of 87% and with a regression R^2^ value higher than 0.99.

### Statistical Analysis

Categorical parameters were expressed as frequencies (%), whereas continuous variables are presented as mean. Data trends were visually and statistically evaluated for normality. We used regression analysis to describe the relationship between categorical variables and between continuous variables with the main outcome. Two-way Student’s t test was then used to detect the difference between Ct values across symptomatic and asymptomatic individuals. Statistical analysis was performed using STATA statistical computer package (StataCorp. 2013. Stata Statistical Software).

### Ethical Approval

The protocol and manuscript for this study were reviewed and approved by the National COVID-19 Research and Ethics Committee in Bahrain (Approval code: CRT-COVID2020-004). This committee has been jointly established by the Ministry of Health and Bahrain Defense Force Hospital research and ethical committees in response to the pandemic, to facilitate and monitor COVID-19 research in Bahrain. All methods and retrospective analysis of data was approved by the National COVID-19 Research and Ethics Committee, and carried out in accordance with the local guideline and ethical guidelines of the Declaration of Helsinki 1975. Informed consent was waived by the National COVID-19 Research and Ethical Committee for this study due to its retrospective and observational nature and the absence of any patient identifying information

## Results

Of the 306 patients, there were 303 who had recovered, whilst 2 deaths and 1 active stable case were excluded from the analysis.

The descriptive summary of the patients is shown in Table 1. The sample of patients included cases who arrived in Bahrain from other countries, all travelers being tested upon arrival, and included cases of local spread. Initially, gender distribution of COVID-19 cases in Bahrain was equal; however, as the infection started to spread within the community of labor workers, who are mainly males, the ratio of males to females and the ratio of local cases to cases coming from other countries increased. Symptomatic patients were 12.2% of the sample, which corresponded to a similar percentage of the symptomatic patients in the entire population.

**Table 1:** Characteristics of patients in the study

| Variable |  | N | % |
| --- | --- | --- | --- |
| Gender | Male | 215 | 71.0% |
|  | Females | 88 | 29.0% |
| Symptoms on Admission | Asymptomatic | 266 | 87.8% |
|  | Symptomatic | 39 | 12.2% |
| Mode of Transmission | Local | 194 | 64.0% |
|  | Travel | 109 | 36.0% |
| Nationality | Bahraini | 177 | 58.4% |
|  | Indian | 89 | 29.4% |
|  | Bangladeshi | 19 | 6.3% |
|  | Other | 18 | 5.9% |
| Category of Ct values | Low : Ct<35 | 271 | 89.4% |
|  | High : Ct>=35 | 32 | 10.6% |
| Variable | N | Mean | Range |
| Age (years) | 303 | 37.1 ± 15.1 | 0 to 87 |
| Ct value on Admission | 303 | 27.5 ± 5.1 | 14.8 to 39 |
| Length of Stay (days)<br>(Time to viral clearance in Days) | 303 | 11.3 ± 5.07 | 4 to 32 |

Ct values showed a statistically significant negative correlation of 0.28 with the length of hospital stay and time to viral clearance as seen in Figure 1 showing the Lowess smoothing curve of hospital stay. Higher Ct values on admission predicted shorter hospital stays and shorter time to clear the virus. The regression analysis showed similar results when using a univariate analysis and a multivariate analysis adjusted for age and gender. Ct value had a significant inverse correlation with the length of stay, 10 points higher on the Ct value correlating with a shorter hospital stay of 2.8 days.

**Figure 1:**
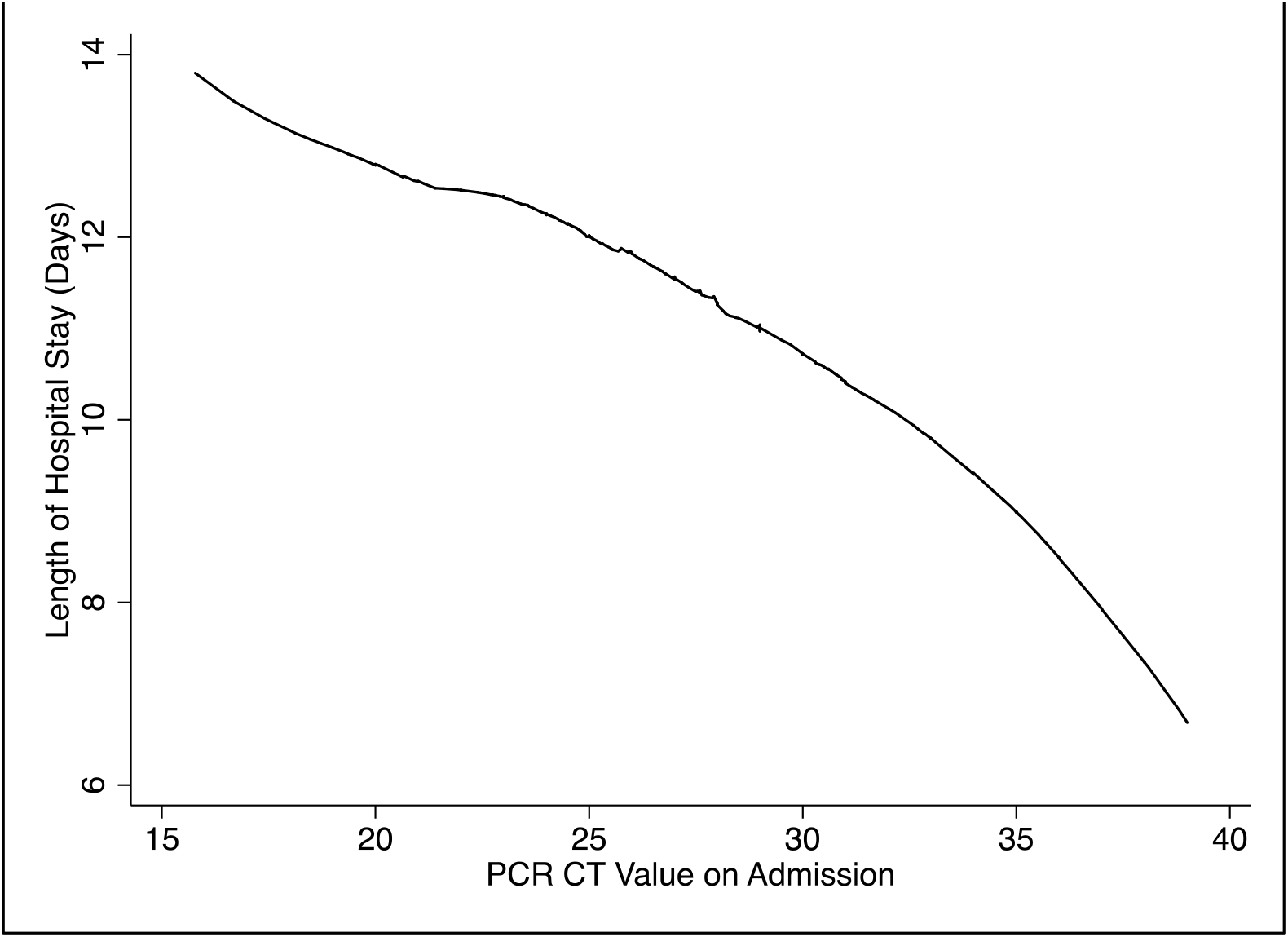
Lowess smoothing curve plotting length of stay (days) vs qPCR Ct value on admission

Patients with high Ct values defined as ≥35 had shorter hospital stays, by 4.3 days, when compared to patients with Ct values <35 on admission.

The regression analysis for Length of Stay using Ct value as a covariate is shown in Table 2. Analysis on symptomatic individuals showed that symptomatic individuals have a lower mean Ct value when compared to asymptomatic individuals. Asymptomatic individuals had a mean Ct value of 27.8. Symptomatic individuals had a mean Ct value of 25.5. A Two sample T-test showed a difference of 2.58 units in the mean Ct value between symptomatic and asymptomatic patients (Table 3).

**Table 2:** Regression analysis for length of stay using Ct value as a covariate

| Multivariate regression output, Adjusted for Age and Gender<br>Outcome: Length of Stay in Days |  |  |  |  |
| --- | --- | --- | --- | --- |
| Continuous covariate | Coefficient | Standard Error | P value | 95% Confidence Interval |
| Ct value | -0.28 | .056 | <0.001 | -0.39 to -0.17 |
| Multivariate regression output, Adjusted for Age and Gender<br>Outcome: Length of Stay in Days |  |  |  |  |
| Categorical covariate | Coefficient | Standard Error | P value | 95% Confidence Interval |
| Low Ct value (<35) | - | - | - |  |
| High Ct value (≥35) | -3.9 | 0.94 | <0.001 | -5.75 to -2.05 |

**Table 3:** Two sample t test comparing mean Ct values between symptomatic and asymptomatic individuals

| Status | Mean Ct value | Standard Error | 95% Confidence Interval |
| --- | --- | --- | --- |
| Asymptomatic | 27.8 | 0.31 | 27.2 to 28.4 |
| Symptomatic | 25.5 | 0.82 | 23.9 to 27.8 |
| Difference | 2.3 | 0.88 | 0.6 to 4.0 |
| P Value |  |  | 0.01 |

## Discussion

This study has shown that the higher the Ct values, the more rapid the viral clearance, leading to a shorter length of stay in hospital. A Ct value >35 predicted a shorter hospital stay by 4.3 days, and a 10 unit increase in Ct value was associated with a shortened time to viral clearance of 2.8 days. Ct values have been reported as an indirect assessment of the viral load ^6, 7^, with higher Ct values having a lower viral load, and it has been shown that those patients with more severe disease have lower Ct values ^7^. Higher Ct values indicate an early infection or a recovering infection. Whilst Ct values can be used to predict length of hospital stay for stable cases, the Ct value cannot, however, predict outcomes. Outcomes are influenced by many additional predictors, most notably comorbidities and severity of disease ^9^. The presence of age greater than 60 and comorbidities including diabetes, hypertension, obesity and respiratory disease may offset a high Ct values and further studies to predict outcomes based on Ct value taking into account adverse risk factors of disease severity are needed.

In this study, symptomatic individuals had lower Ct values whilst asymptomatic individuals had higher Ct values, suggesting that the lower Ct values reflects a more severe presentation. However, asymptomatic individuals still had a low mean Ct values (27.8) indicating a significant viral load, suggesting that asymptomatic individuals can still transmit the disease ^10^. These results are in accord with those of Liu et al who reported that the mean viral load of severe cases was around 60 times higher than that of mild cases ^7^. La Scola et. al have reported using cell culture system that patients with Ct values >= 34 do not shed infectious virions, and hence are not infectious and can be discharged from hospitals or care centers^11^.

Bahrain has pursued a comprehensive testing program with all travelers arriving in the country being tested, mobile screening services, random screening in high risk communities and extensive contact tracing. This strategy has resulted in detecting many asymptomatic patients, in contrast to other countries where the response has been directed solely towards symptomatic patients, and has generated data that is critical for production of accurate prediction models ^12^. Limitations of this study include that it was cross sectional and that the number of symptomatic cases were few in number. The number of patients with multiple comorbidities were few and therefore a model to take into account the protective effect of a high Ct value could not be determined.

## Conclusion

In this current study it was evident that Ct value can predict the duration of hospital stay of COVID-19 infected patients, which also suggests that viral clearance will be faster in higher Ct value patients. This data can be implemented in other country to evaluate the patients stay at hospital and also helpful in managing the healthcare staff in successfully handling the COVID-19 infected patients.

## Data Availability

Available upon reasonable request

## DECLARATIONS

### Ethics approval and consent to participate

The study was approved by the National Covid-19 Research and Ethics Committee.

### Consent for publication

Consent for publication was waived by the Ethical committee.

### Availability of data and materials

All the data for this study will be made available upon reasonable request to the corresponding author.

### Author contributions

SA, and OY analyzed the data and wrote the manuscript. AA, OY and SA contributed to study design, collected, analysed, and interpreted data and edited the manuscript. MA supervised data collection, data analysis and edited the manuscript. EMJ data interpretation and the editing of the manuscript. All authors reviewed and approved the final version of the manuscript. Manaf Alqahtani is the guarantor of this work.

